# Cost-effectiveness of a virtual fracture clinic versus traditional in-person fracture clinic care for adults with acute simple fractures: a protocol for a health economic evaluation within the RECITAL trial

**DOI:** 10.64898/2026.06.16.26355732

**Authors:** Rowena Charteris, Adrian C Traeger, Christopher G Maher, Tessa Copp, Kristen Pickles, Min Jiat Teng, Isabella Khoudair, Ben Warnock, Miranda Shaw, Owen Hutchings, Mark Horsley, Ilana N Ackerman, Ray Thomas, Philip Haywood, Joshua R Zadro

**Author notes:** Consumer representative (Ray Thomas). Philip Haywood and Joshua Zadro are joint-senior authors.

## Abstract

**Introduction:** Traditional in-person fracture clinics are often overcrowded and inconvenient for patients. Virtual fracture clinics aim to address some of these concerns by improving the efficiency of the orthopaedic service and reducing unnecessary interventions while maintaining safety and quality of care. The RECITAL trial is a non-inferiority randomised controlled trial comparing follow-up care provided at a virtual fracture clinic for people with acute simple fractures to follow-up care provided at an in-person fracture clinic. This study describes the protocol for an economic evaluation of RECITAL where the primary aim is to investigate the cost-effectiveness of a virtual fracture clinic compared with traditional in-person fracture clinic care from a health system perspective.

**Methods and analysis:** The RECITAL trial recruited 312 participants with acute simple fractures and randomised them to receive follow-up care provided at a virtual fracture clinic or follow-up care provided at an in-person fracture clinic. We will conduct a within-trial analysis from a health system perspective (primary analysis), as well as a health service, patient and societal perspective. The economic evaluation will estimate the difference in the cost of resource inputs on an intention to treat basis used by participants in the two arms of the trial, allowing comparisons to be made between the in-person and virtual fracture clinics. Data for intervention costs and healthcare utilisation will be collected from trial records, hospital electronic medical records and district performance units. The results of the economic evaluation will be expressed in terms of incremental cost per utility weight gained at 12 weeks and will be plotted on a cost-effectiveness plane. Bootstrapping by resampling will be used to estimate 95% confidence intervals around costs and outcomes, and to calculate the confidence intervals around the incremental cost-effectiveness ratio. A cost-effectiveness acceptability curve (CEAC) will be plotted, which will provide information about the probability that an intervention is cost-effective, given the level of a decision maker’s willingness to pay for each additional outcome.

**Ethics and Dissemination:** The trial was approved by the SLHD Ethics Review Committee (RPAH Zone) (X23-0200 and 2023/ETH01038). The findings will be disseminated through a peer-reviewed journal and conference presentations.

Trial registration number

The trial was prospectively registered on the Australian New Zealand Clinical Trials Registry (ANZCTR; 12623000934640)

## INTRODUCTION AND BACKGROUND

Musculoskeletal conditions represent a significant and growing burden on health systems worldwide, driven in part by high rates of emergency department (ED) presentations and rising rates of fractures requiring care in orthopaedic fracture clinics.^1^ In Australia, spending on musculoskeletal disorders has consistently been in the top 4 disease groups each year from 2013-23.^2^ The total expenditure on musculoskeletal disorders within Australian Hospitals alone in 2023-24 was $11 billion.^2^ In 2024-25 musculoskeletal injuries made up 5.3% of the 9 million presentations to Australian emergency departments.^3^ Globally, there were 178 million new fractures in 2019, increasing over 33% since 1990.^4^ This growth places increasing pressure on health systems and has been the catalyst for trials looking to redesign patient pathways to improve efficiency.^5,6^

Traditionally, patients diagnosed with acute fractures have had their follow-up care provided in face-to-face orthopaedic clinics. These clinics are often oversubscribed, inefficient and can be inconvenient for patients.^7^ Virtual Fracture Clinics (VFC) have become increasingly common in the UK, with the aim to reduce patient length of stay in the Emergency Department and to improve efficiency of the orthopaedic service by reducing unnecessary in-person appointments and repeat imaging.^8^ Patients typically referred to virtual fracture clinics are those with acute fractures that do not require immediate orthopaedic admission and can be managed with removable splints. These patients are typically referred from the ED to a virtual fracture clinic for any follow-up care such as splint removal, education on prognosis, reassurance, and advice on activities.^5^

In recent systematic reviews, Virtual Fracture Clinics have been found to improve patient access to care and reduce wait times for an appointment, without increasing the risk of adverse events or re-presentations.^8,9^ Several retrospective observational studies have explored the costs of virtual fracture clinics. These studies have demonstrated financial benefits to the health service,^10-13^ largely attributed to fewer patients being seen at in-person clinics, improved staffing efficiencies, and fewer unnecessary appointments and imaging.^14^ However, no trial-based economic evaluation exists for this model of care.

To the best of our knowledge, the RECITAL trial is the first randomised controlled trial assessing the safety and clinical effectiveness of a virtual fracture clinic.^15^ Trial-based economic evaluations offer important advantages over those derived from observational data, including stronger causal inference through randomisation, prospective collection of resource use and outcomes, and reduced risk of selection bias. The primary objective of this health economic evaluation is to investigate the cost-effectiveness of a virtual fracture clinic compared with traditional in-person fracture clinic care from a health system perspective. This will be achieved by comparing cost per additional EQ-5D utility weight difference gained at 12 weeks between groups. The secondary objective is to investigate the cost-effectiveness of a virtual fracture clinic from the health service (Sydney Local Health District) perspective, the patient’s perspective and a societal perspective.

## METHODS

### Study design

We will conduct an economic evaluation of the RECITAL trial; a non-inferiority randomised controlled trial comparing follow-up care provided at a virtual fracture clinic for people with acute simple fractures to follow-up care provided at an in-person fracture clinic. A within-trial analysis will be conducted from a health service, health system, patient and societal perspective, with the primary focus being on cost-effectiveness from a health system perspective. The economic assessment method will be reported according to the Consolidated Health Economic Evaluation Reporting Standards 2022 (CHEERS 2022).^16,17^ The RECITAL trial is briefly described below. Further details can be found in the published trial protocol.^15^

The RECITAL trial is a prospective two-arm, parallel group non-inferiority randomised controlled trial conducted within Sydney Local Health District, titled “Effects of virtual fractuRE Clinic care compared with In-person fracture clinic care on physical function in people with simple fractures: a non-inferiority randomised TriAL (RECITAL)”. The trial was prospectively registered on the Australian New Zealand Clinical Trials Registry (ANZCTR; 12623000934640) and was approved by the SLHD Ethics Review Committee (RPAH Zone) (X23-0200 and 2023/ETH01038). A consumer representative was involved in the development and conduct of the trial and will be involved in this economic evaluation.

### Participants and recruitment

The RECITAL trial recruited 312 individuals 18 years or older who were referred to the virtual fracture clinic at Sydney Local Health District Virtual Hospital (Sydney Virtual, formerly RPA Virtual) for follow up care of an acute (<6 weeks), simple fracture. Participants were deemed appropriate for remotely managed care by a multidisciplinary team, including an orthopaedic doctor. Patients were excluded if they had a complex or significantly displaced fracture, including pathological, open, unstable or spinal fractures; required a cast or surgical management; had neurovascular concerns; or had a condition not managed by the Royal Prince Alfred Hospital (RPA) Orthopaedic Department. Patients who did not provide informed consent or were unable to attend a clinic within the recommended time were also excluded. Recruitment began in November 2023 and was completed in March 2025. Participants were followed up to 12 weeks post-randomisation. Collection of follow-up data concluded in June 2025.

Characteristics of the participants at baseline can be found in **Table 1**. below.

**Table 1.**

| **Characteristic** | **Virtual**<br>**(N=156)** | **In-person**<br>**(N=156)** |
| --- | --- | --- |
| **Median age (IQR) – yr** | 42 (29-54) | 40 (30-55) |
| **Gender – no. (%)** |  |  |
| Female | 97 (62.2%) | 89 (57.1%) |
| Male | 59 (37.8%) | 67 (42.9%) |
| **Region of birth** |  |  |
| Australia & New Zealand | 114 (73.1%) | 105 (67.3%) |
| Western/Northern Europe | 16 (10.3%) | 20 (12.8%) |
| Southern/Eastern Europe | 8 (5.1%) | 6 (3.8%) |
| Asia | 7 (4.5%) | 11 (7.1%) |
| North America | 4 (2.6%) | 6 (3.8%) |
| Africa and Middle East | 4 (2.6%) | 5 (3.2%) |
| South/Central America & Caribbean | 3 (1.9%) | 3 (1.9%) |
| **Highest level of education** |  |  |
| Doctorate or Master's Degree | 27 (17.3%) | 30 (19.2%) |
| Graduate Certificate or Diploma | 12 (7.7%) | 9 (5.8%) |
| Bachelor Degree with Honours | 13 (8.3%) | 16 (10.3%) |
| Bachelor Degree or Associate Degree | 41 (26.3%) | 43 (27.6%) |
| Diploma (incl. advanced or associate) | 12 (7.7%) | 15 (9.6%) |
| Certificate I-IV or Advanced | 18 (11.5%) | 20 (12.8%) |
| Secondary Education only or not stated | 33 (21.2%) | 23 (14.7%) |
| **Fracture diagnosis** |  |  |
| Metatarsal | 38 (24.4%) | 41 (26.3%) |
| Toe | 41 (26.3%) | 34 (21.8%) |
| Stable ankle fracture | 18 (11.5%) | 33 (21.2%) |
| Elbow | 27 (17.3%) | 21 (13.5%) |
| Tarsal avulsion | 20 (12.8%) | 18 (11.5%) |
| AC joint injury | 2 (1.3%) | 4 (2.6%) |
| Triquetral | 5 (3.2%) | 1 (0.6%) |
| Clavicle | 2 (1.3%) | 2 (1.3%) |
| Other | 8 (5.1%) | 10 (6.4%) |

### Randomisation

Participants were randomly assigned to receive their follow-up care in a virtual fracture clinic (intervention group) or an in-person fracture clinic (control group). Randomisation was computer generated within REDCap using random blocks of 4, 6, 8 and 10. Study assessors and statistician were blinded to group allocation.

### Interventions

The virtual fracture clinic group received their follow-up care via phone or video calls with a physiotherapist and were emailed a standard fracture management fact sheet. The in-person care group received follow-up care at the in-person fracture clinic with an orthopaedic doctor.

### Trial outcomes

Trial outcomes were collected at baseline and at 6 and 12 weeks post-randomisation. The primary outcome was self-reported physical function measured with the Patient-Specific Functional Scale (PSFS) at 12 weeks. The secondary outcomes included the PSFS at 6 weeks, pain (assessed via the Pain Numerical Rating Scale, NRS) at 6 and 12 weeks, patient experience (assessed via the Generic Short Patient Experience Questionnaire) at 6 weeks, health care utilisation at 12 weeks, medication use at 6 weeks, health-related quality of life (assessed via the EuroQol five-dimension five-level questionnaire, EQ-5D-5L) at 12 weeks, and adverse events at 6 and 12 weeks.

### Economic data collection

For the economic evaluation, we will use the following data from the RECITAL trial: patient demographics (age, total income, education levels, and health literacy levels), fracture site, health service resource utilisation from the Sydney Virtual and in-person clinics (e.g. workforce, infrastructure and technology, and consumables), radiology scans, ED re-presentations, other healthcare visits (e.g. GP or physiotherapy), travel costs and loss of productivity.

Information will be gathered from the patient’s hospital medical records, the hospital’s Targeted Activity and Reporting System (STARS App) Dashboard, the SLHD or Sydney Virtual performance units, and through the REDCap survey used to collect study data at baseline, 6-weeks and 12-weeks post randomisation in RECITAL.

The EQ-5D-5L survey will be used to generate utility weights. EQ-5D-5L responses will be transformed by the national Australian value set for the EQ-5D-5L into utility weights.^18,19^

The gain in outcomes at an individual level will be estimated using the differences in the utility weights and the PFSF between the baseline and follow-up surveys.

### Measurement and valuation of resources

Costs will be calculated for each individual in the study, by multiplying the relevant resource use by the corresponding valuation. Costs for resource inputs will largely be derived from local and national sources and estimated in line with best practice. Primary research using established accounting methods may also be required to estimate unit costs. Costs will be standardised to current prices where possible.

Measurement and valuation of resource use can be found in **Table 2**. This data will be collected using trial records and patient reported outcome measures and will be cross-checked with patient electronic medical records to ensure accuracy and completeness.

**Table 2.**
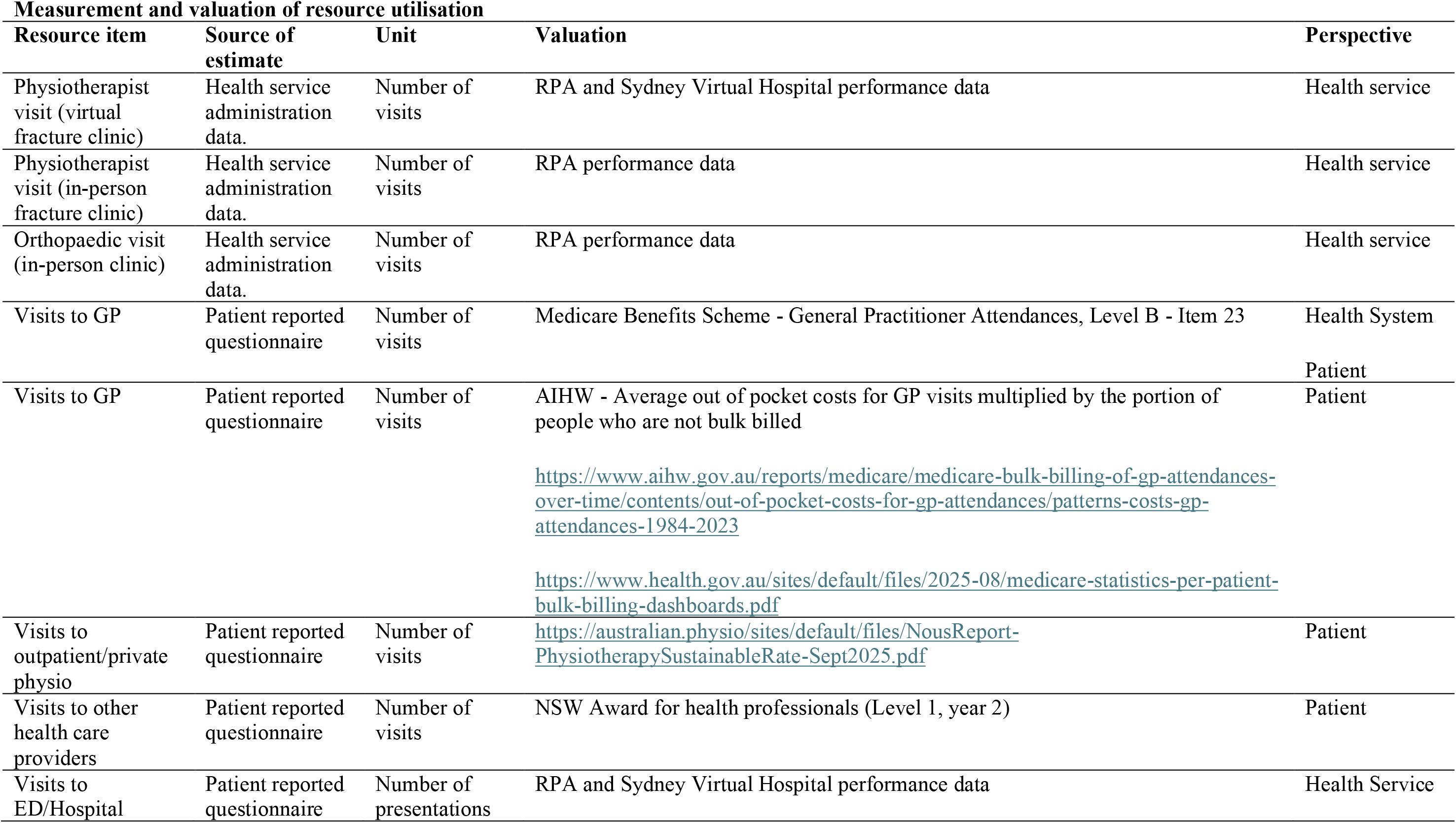
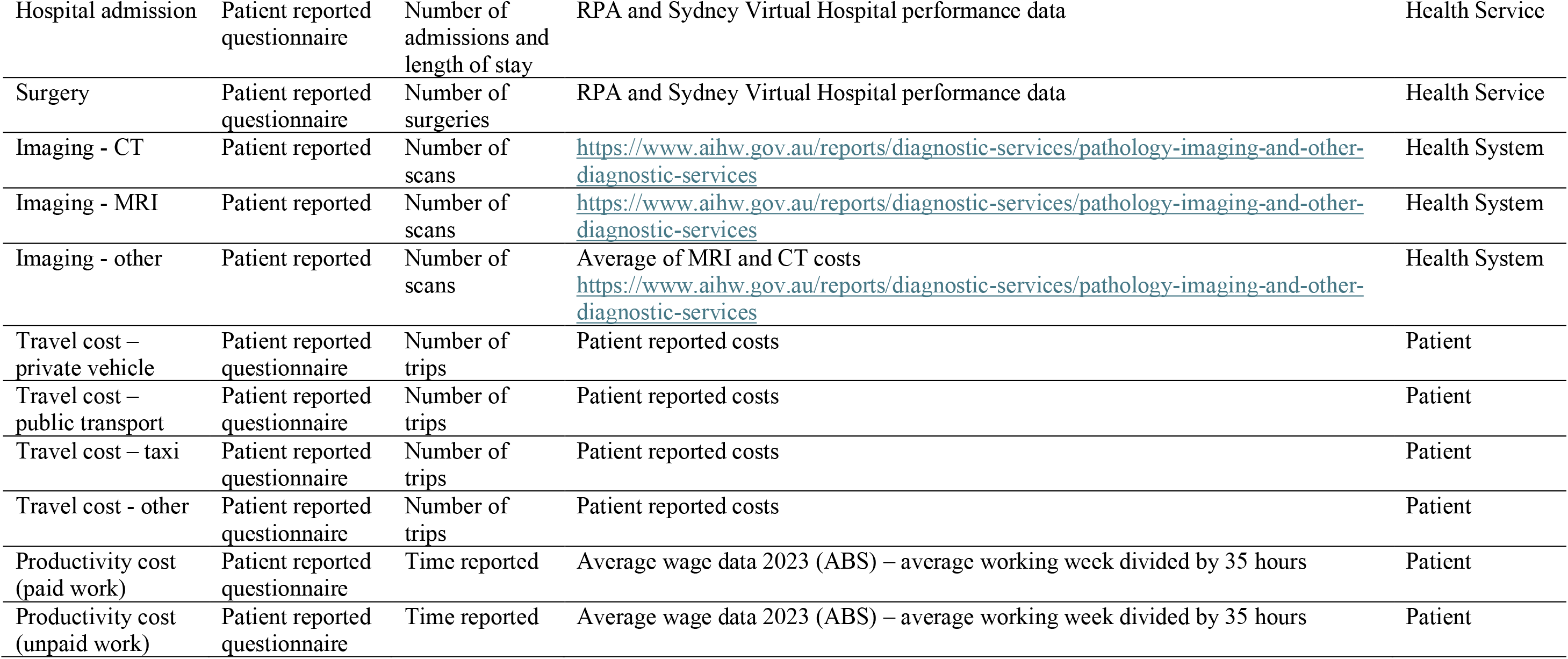

Costs will be valued in Australian dollars using relevant state and national standards for the financial year 2023/24 with indexation measured using the Australian Institute of Health and Welfare deflator, Australian Bureau of Statistics Consumer Price Index (CPI) or wages indexation as required. Discounting will not be required as the study duration is less than one year.

### Economic evaluation analysis

For the primary analysis, a cost-effectiveness analysis will be performed from the perspective of the health system. This perspective was chosen because it is the most useful for decision-makers in the context of adopting new models of care by the funders and managers of hospital care. The patient’s perspective will be included in the sensitivity analysis and may give important contextual information to those decision-makers.

The RECITAL trial models of care ran over a 12-week period, which was decided because the majority of uncomplicated fractures will have had clinical union by 12 weeks and patients will have been discharged.^20^ Any between group differences arising after this point were thought to be unrelated to the model of care. As such, the time horizon of the economic evaluation will be 12 weeks.

Data analysis will be conducted using Stata (StataCorp. 2025. Stata Statistical Software: Release 16. College Station, TX: StataCorp LLC). Descriptive statistics will be used to summarise the data.

The economic evaluation will estimate the difference in the cost of resource inputs on an intention to treat basis used by participants in the two arms of the trial, allowing comparisons to be made between the in-person and virtual fracture clinics.

We will conduct a cost-effectiveness analysis to estimate the incremental cost per utility weight difference at 12 weeks defined as: [cost of the virtual care - cost of in-person care]/ [effectiveness of the virtual care – effectiveness of in-person care]. The results of the economic evaluation will be expressed in terms of incremental cost per utility weight gained at 12 weeks and will be plotted on a cost-effectiveness plane. Multiple imputation methods will be used to impute missing data and avoid biases associated with complete case analysis as documented in the RECITAL trial statistical analysis plan.^21^

Bootstrapping by resampling will be used to estimate 95% confidence intervals around costs and outcomes, and to calculate the confidence intervals around the incremental cost-effectiveness ratio. Seemingly unrelated regression will be used to account for the potential correlation between costs and outcomes.

A cost-effectiveness acceptability curve (CEAC) will be plotted, which will provide information about the probability that an intervention is cost-effective, given the level of a decision maker’s willingness to pay for each additional outcome.

Subgroup analysis will be carried out to assess the heterogeneity and the equity impact of the interventions between participants with upper vs lower limb fractures, age, total income, participant education levels, and health literacy levels.

## SENSITIVITY ANALYSES

Secondary analysis will be conducted from a health services perspective (Sydney Local Health District) and patient’s perspective, as well as a societal perspective by combining the health system and patient perspectives. One-way sensitivity analyses will be conducted around key cost variables, including the average costs of providing virtual vs in-person fracture care beyond the trial period (e.g., 2024 and 2025) and variations to estimates for attributed costs to Sydney Virtual and RPA hospital, outpatient healthcare provider costs and patient productivity. A probabilistic sensitivity analysis will be conducted to estimate the joint uncertainty in all parameters.

## Data Availability

All data produced in the present study are available upon reasonable request to the authors

## Data Availability

All data produced in the present study are available upon reasonable request to the authors

